# Towards a Robust cell-free DNA Isolation Protocol for NGS Applications in a Clinical Molecular Diagnostics Setting

**DOI:** 10.64898/2026.06.15.26355337

**Authors:** Melina Apweiler, Julian Broche, Marion Loitz, Luise Hackenbruch, Stephan Ossowski, Christopher Schroeder, Kristopher J Schmit

## Abstract

Cell-free DNA (cfDNA), released from apoptotic and necrotic cells into body fluids, represents a non-invasive source of genetic information for disease prediction, diagnosis, and monitoring. However, its low physiological abundance makes cfDNA highly susceptible to pre-analytical influences. In particular, genomic DNA (gDNA) released from lysed white blood cells (WBCs) can contaminate plasma and compromise downstream cfDNA analyses. This study evaluated the impact of different blood collection tubes and isolation methods on cfDNA stability and yield. Blood samples from 13 healthy donors were collected using cfDNA-stabilizing tubes (Cell-Free DNA BCT®, Streck; S-Monovette® cfDNA Exact, Sarstedt) and stored at room temperature for 1, 5, or 10 days before plasma isolation. CfDNA was extracted using either a magnetic bead-based method or a silica column-based approach. DNA quantity and quality were assessed by fluorometric quantification, automated fragment analysis, and gene-specific quantitative PCR. Streck-based workflows maintained stable cfDNA yields and characteristic mononucleosomal fragmentation profiles across all storage times. In contrast, Sarstedt tubes showed reduced cfDNA concentrations after 5 days and a pronounced increase at *10 Days*, accompanied by high–molecular weight DNA patterns consistent with WBC lysis. These trends were largely independent of the extraction method. Overall, the results demonstrate that blood collection tube chemistry critically influences cfDNA integrity during delayed processing. Streck tubes, particularly when combined with QIAamp®, provided the most robust and reproducible workflow for routine molecular diagnostics, whereas Sarstedt tubes produced physiologically implausible results after extended storage.

**Key Points:**

- Extended standing times of BCTs prior to plasma isolation have the greatest impact on cfDNA integrity
- Streck BCTs outperform Sarstedt BCTs for cfDNA yield and quality
- The cfDNA isolation method only plays a secondary role

## Introduction

Liquid biopsies (LBs) have emerged as a promising non-invasive alternative to tissue biopsies in molecular diagnostics. A variety of bodily fluids including saliva, urine, cerebrospinal fluid, amniotic fluid, and particularly blood serve as accessible matrices for the detection and longitudinal monitoring of circulating biomarkers in health and disease. Among these, circulating cell-free DNA (cfDNA), released predominantly during apoptosis, has gained considerable attention as a source of genetic and epigenetic information. Apoptotic fragmentation typically yields mononucleosomal cfDNA fragments of approximately 160–180bp (1–3).

CfDNA is increasingly investigated as a biomarker across diverse clinical applications, including non-invasive prenatal testing (NIPT), as well as early detection and monitoring in oncology and neurodegenerative diseases (4–6). In Germany, NIPT is clinically approved for the detection of fetal aneuploidies, via quantification of fetal cfDNA from the placenta in maternal blood (5) . In oncology, cfDNA, more specifically circulating tumor DNA (ctDNA), can be used for early cancer detection, molecular tumor characterization, therapy monitoring, or minimal residual disease detection (6,7). Given that these applications traditionally rely on invasive tissue biopsies, and considering cancer as the second leading cause of mortality, cfDNA-based diagnostics hold significant potential to improve clinical outcomes (6,8).

Downstream cfDNA analyses typically involves polymerase chain reaction (PCR)-based or next-generation sequencing (NGS)-based approaches, such as digital PCR and whole-genome sequencing (6,7). The performance of these methods with respect to sequencing depth, coverage, and variant detection rely greatly on cfDNA purity and integrity. In healthy individuals endogenous cfDNA concentrations in blood plasma are low (< 10 ng/mL), whereas in malignancies plasma cfDNA levels commonly surpass 20 ng/mL and can even be beyond 1,000 ng/mL depending on tumor type and stage (7,9–11). A major analytical challenge lies in the low abundance of target-derived cfDNA. In NIPT, fetal cfDNA constitutes only a minor fraction of total maternal cfDNA; likewise neuron- or glia-derived plasma cfDNA occurs at very low abundance in neurological disorders, limiting detection sensitivity (5,12). This is largely due to the predominance of cfDNA originating from hematopoietic cells (12). Additionally, cfDNA can be diluted or contaminated by genomic DNA (gDNA) released from lysed blood cells. Such gDNA contamination is exacerbated by suboptimal transport conditions, particularly temperature fluctuations and most importantly prolonged storage of whole blood, potentially compromising downstream analyses and leading to inaccurate results (11,13–15).

To mitigate these pre-analytical challenges, cfDNA-stabilizing blood collection tubes (BCTs) have been developed to preserve cfDNA integrity. Compared to conventional EDTA tubes, these specialized BCTs have demonstrated enhanced cfDNA stability, likely due to the inclusion of anti-lysis reagents (16–18). Additionally, multiple cfDNA isolation kits have become available, based on different isolation principles, among which the most common are magnetic-bead-based or silica-column-based. Both, BCT type and cfDNA isolation kit can influence cfDNA yield, purity and integrity.

Accordingly, a systematic evaluation of these factors was conducted in the Department of Molecular Diagnostics at the Institute of Medical Genetics and Applied Genomics, Tübingen. This randomized mixed factorial study aimed to assess the robustness of current workflows and to optimize standard operating procedures for the generation of high-quality cfDNA suitable for downstream NGS applications in routine molecular diagnostics.

## Material and Methods

### Ethics and volunteer recruitment

Study was conducted based on principles of the Declaration of Helsinki and approval obtained from the Institutional Ethics Committee of the University Hospital Tübingen (No. 173/2011BO2 and 186/2017BO1). Venous blood was collected from 13 healthy adult volunteers (sex-balanced), recruited at the Fertility Counseling Unit, Department for Women’s Health, and the Institute of Medical Genetics and Applied Genomics, Tübingen University Hospital (Tübingen, Germany) (**Supplementary Table 1**). Detailed inclusion and exclusion criteria are listed in **Supplementary Table 2**.

### Blood Collection

Blood was collected into two different cfDNA-stabilizing blood collection tubes (BCTs) (**Figure 1**): Cell-Free DNA BCT® (cat. no. 218997; Streck LLC, La Vista, NE, USA) and S-Monovette® cfDNA Exact (cat. no. 01.2040.001; SARSTEDT AG & Co. KG, Nümbrecht, Germany), hereafter referred to as *Streck* and *Sarstedt*, respectively. For 11 out of 13 volunteers, two of each cfDNA BCT à 5 mL blood were collected and assigned to either the *short-standing time* group (6/11 volunteers) or the *long-standing time* group (5/11 volunteers) (**Supplementary Figure 1**, **Supplementary Table 1**). For each of these volunteers, one of both BCT types was processed after 1 day (D01). The second tube was processed either after 5 days (D05; short standing time group) or at 10 Days (D10; long standing time group). For two volunteers, only one tube per BCT type was collected; these samples were exclusively processed at 10 Days (**Supplementary Table 1**).

**Figure 1.**
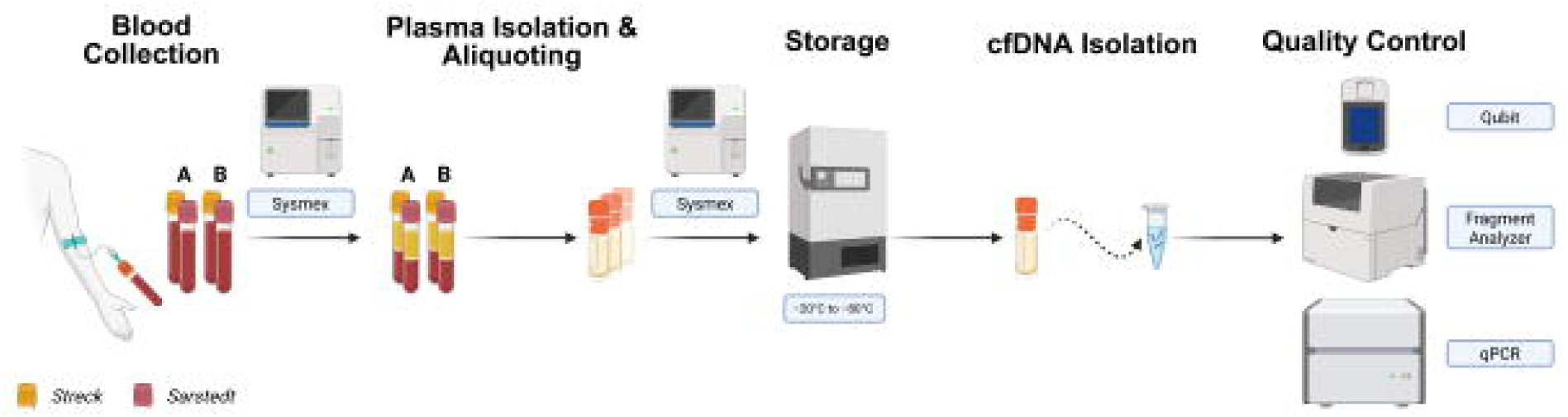
Experimental workflow of the study. Per patient, venous blood was collected into two Cell-Free DNA BCT® tubes (Streck; yellow cap) and two S-Monovette® cfDNA Exact tubes (Sarstedt; pink cap). Samples were divided into two experimental sets. Set A tubes were processed after 24 h (1 day) of standing time. Set B were processed either after 120 h (5 days) or 240 h (10 days). Prior and post plasma isolation all samples were assessed using a Sysmex hematology analyzer. The isolated plasma was aliquoted in cryotubes and at –20 °C for short-term (–80 °C for long-term) storage. Cell-free DNA (cfDNA) was isolated using either a magnetic bead-based method (MagMAX™ Cell-Free DNA Isolation Kit) or a silica column-based method (QIAamp MinElute ccfDNA Kit). Downstream analyses for quality- and quantity control included fluorometric DNA quantification using Qubit, fragment size distribution analysis using the TapeStation system, and gene-specific DNA quantification by quantitative PCR (qPCR).

Blood collection was done in accordance with the manufacturers’ instructions. Blood collection and all downstream processing steps were performed in the same building under stable ambient conditions.

### Hematological Analyses and Plasma Isolation

BCTs were stored upright at room temperature (RT) until their respective day of processing. To monitor cellular integrity and hemolysis over time, complete blood count (CBC) was performed on a Sysmex XP-300 hematology analyzer (Sysmex Corporation, Kobe, Japan) according to manufacturer’s instructions, prior and post plasma isolation (**Figure 1**). All measured values are provided in **Supplementary Table 3**.

### Plasma isolation

Plasma was isolated using a two-step centrifugation protocol (**Figure 1**). BCTs were centrifuged at 2,000 × *g* for 10 min at RT, and resulting supernatants transferred into 15 mL conical tubes (cat# 188 271-N; Greiner Bio-One, Kremsmünster, Austria), which were centrifuged again at identical settings. The cleared plasma was then aliquoted in cryotubes (cat# 65-7641; Azenta, Burlington, USA) and stored at −20 °C until further processing (**Figure 1**).

### (cf)DNA Isolation

→ see *Figure 1*

Plasma preparation

A starting plasma volume of 1.3–1.5 mL was thawed at 37 °C for 10 min on a Thermomixer (Eppendorf SE, Hamburg, Germany). The total plasma volume was transferred to a fresh 15 mL conical tube and adjusted to a total volume of 2 mL with 1× Phosphate Buffered Saline (PBS) (cat# 10010-023; Thermo Fisher Scientific, Waltham, USA).

Magnetic Bead-Based Isolation (MagMAX™Cell-free DNA Isolation Kit)

This protocol was performed according to manufacturer’s instructions. In short, thawed sample were incubated with Proteinase K (cat# AM2548; Thermo Fisher Scientific, Waltham, USA), and 20% sodium dodecyl sulfate (SDS) (cat# AM9820; Thermo Fisher Scientific, Waltham, USA) at 60 °C for 20 min at 400 rpm, followed by immediate cooling down on ice for 5 min. Then, Cell-Free Lysis/ Binding Solution (cat# A33600; Thermo Fisher Scientific, Waltham, USA), and Magnetic Bead Solution (cat# 37002D; Thermo Fisher Scientific, Waltham, USA) were added, incubated on a Rotator (Antylia Scientific, Vernon Hills, USA) at RT for 10 min. Samples were briefly spun down for 30 sec at 200 × *g*, and placed on a magnetic stand until solution cleared. Supernatants were discarded, and beads were washed twice, respectively, first with Wash Solution (cat# A33601; Thermo Fisher Scientific, Waltham, USA) and next with 80% ethanol in 1.5 mL LoBind tubes (cat# 022431021; Eppendorf SE, Hamburg, Germany). (cf)DNA was eluted in 54 µL Elution Solution (cat# A33602; Thermo Fisher Scientific, Waltham, USA) and incubated for 5 min at 2,000 rpm at RT. Clear eluates were transferred into new 1.5 mL LoBind tubes and stored at –20 °C until further analysis.

Silica Column-Based Isolation (QIAamp® MinElute ccfDNA Kit)

This protocol was performed according to manufacturer’s instructions. In short, proteinase K, magnetic beads and Bead Binding Buffer were added to the plasma sample and incubated at RT for 10 min on a Rotator. After the following magnetic separation, bead pellets were resuspended in 200 µL Bead Elution Buffer, transferred into Bead Elution Tubes and incubated for 5 min at 300 rpm at RT. After the next magnetic separation, the clear eluates were transferred into new Bead Elution Tubes and mixed with 300 µL ACB Buffer and were then loaded onto QIAamp UCP MinElute columns and centrifuged for 1 min at 6,000 × *g*. Columns were placed into fresh 2 mL collection tubes and centrifuged for 3 min at 20,000 × *g* to dry the membranes completely. Each column was transferred into a fresh 1.5 mL Elution Tube and incubated with open lids at 56 °C for 3 min. Finally, cfDNA was eluted in 60 µL Ultra-clean water. After 1 min incubation at RT with closed lids, columns were centrifuged at 20,000 × *g* for 1 min. This step was repeated to maximize cfDNA yield. CfDNA eluates were stored at –20 °C until further analysis.

### Quality Control of Isolated (cf)DNA

→ see *Figure 1*

Fluorometric DNA Quantification

Absolute DNA concentrations were determined by Qubit™ dsDNA High Sensitivity (HS) Assay Kit (cat# Q32851; Thermo Fisher Scientific, Waltham, USA), according to manufacturers’ protocol. Measurements were performed on a Qubit™ 3.0 Fluorometer (Thermo Fisher Scientific, Waltham, USA). For cfDNA samples below the limit of detection, 4 µL of eluted cfDNA were mixed with 196 µL of working solution prior to measurement.

Fragment Size Distribution Analysis

DNA fragment size distributions and integrity were assessed via electrophoresis-based measurements on an Agilent 4200 TapeStation (Agilent Technologies, Santa Clara, USA) using the Cell-Free DNA ScreenTape assay (cat# 5067-5630; Agilent Technologies, Santa Clara, USA) according to manufacturer’s instructions.

Gene-Specific Quantification

Quantitative PCR (qPCR) was performed to quantify single-copy genes *SLC6A3* and *MSTN*. Primers for *SLC6A3* (98 bp amplicon) and *MSTN* (88 bp amplicon) (Integrated DNA Technologies, Coralville, USA) were used (**Supplementary Table 4A**). The assay was performed for total reaction volume of 10 µL with 5 µL 2× QuantiTect SYBR Green PCR Master Mix (cat# 204343; Qiagen, Hilden, Germany), 0.3 µL of either forward or reverse primers (1:10 dilution), and 1 µL template cfDNA. Reactions were run in triplicates, including a No Template Control (NTC) for each primer pair. Reactions were set up in 384-well plates (cat# RT-PL384-LCW; Eurogentec, Seraiyng, Belgium), sealed, centrifuged for 2 min at 200 × *g*, and analyzed on Roche LightCycler® 480 (Roche, Basel, Switzerland), using the program as seen in **Supplementary Table 4B**.

To assess qPCR efficiency and perform a total quantification analysis, a dilution series of 100%, 20% and 4% of an additional cfDNA sample with known concentration (0.292 ng/µL) was added. The group *StreMax* at D01 was used as reference group for data analysis.Data Analysis and Statistics

Preliminary processing and normalization of raw data was performed in MS Excel. For non-parametric statistical analysis and visualization, Python (version 3.12.11) was used. Pairwise comparison between Streck and Sarstedt samples were assessed using Mann-Whitney U test. For all other comparisons, Kruskal-Wallis test was applied, followed by Dunn’s post-hoc with Benjamini-Hochberg to correct for multiple comparisons. Statistical significance interpreted as: ns, p > 0.05; *, p ≤ 0.05; **, p ≤ 0.01; ***, p ≤ 0.001. Results were reported as median values with corresponding interquartile ranges (IQR). Additionally, relative intragroup changes for *5 Days* and *10 Days* to *1 Day* were reported as Δ.

## Results

### Sarstedt BCT plasma has lower residual cellular content than Streck

CfDNA is the collection of fragmented DNA originating from physiological or disease triggered cell death. Among the most important contaminations to avoid when working with cfDNA are hemolysis and cellular genomic contamination after blood withdrawal (15). Specialized stabilizing BCTs and varying standing times are two crucial factors affecting the degree of such contamination. In this study, six essential hematological parameters (red blood cells (RBCs), white blood cells (WBCs), hemoglobin (HGB), hematocrit (HCT), platelets (PLTs; ×10^3^/µL), and plateletcrit (PCT)) were assessed prior (**Supplementary Figure 2, Supplementary Table 4**) and post (**Figure 2**) plasma isolation from either Streck or Sarstedt BCTs after one, 5- and 10-days standing time.

**Figure 2.**
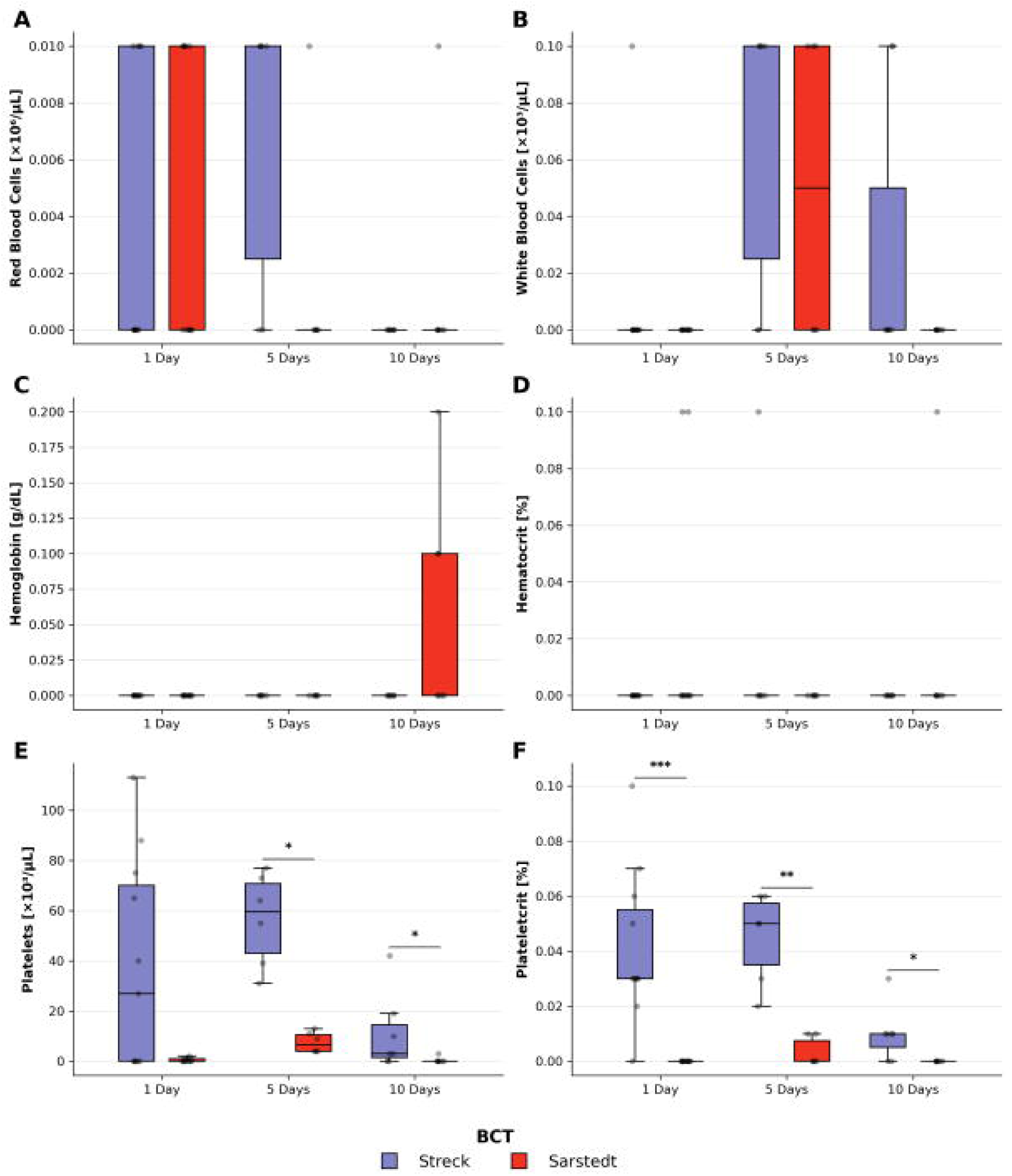
Hematological assessment of plasma isolated from Streck and Sarstedt blood collection tubes using Sysmex hematology analyzer. Boxplots illustrating the temporal changes in six hematological parameters between Streck and Sarstedt BCTs. These six parameters are A) red blood cells (RBCs) and B) white blood cells (WBCs) counts, C) hemoglobin (HGB) concentrations, D) hematocrit (HCT) ratios, E) platelet (PLT) counts, and F) plateletcrit (PCT) ratios. Statistics were performed using Mann-Whitney U testing. *, p ≤ 0.05; **, p ≤ 0.01; ***, p ≤ 0.001

Prior to plasma isolation, all parameters decreased, oftentimes significantly (not shown; **Supplementary Table 7**), over time aside from HGB, which was practically not detected (**Supplementary Figure 2**). Notable was the significantly (p = 0.0227) higher RBC counts in Streck as in Sarstedt BCTs and vice-versa, while not statistically significant, for WBC counts (p = 0.0856) (**Supplementary Figure 2**, **Supplementary Table 6**). After plasma isolation, RBC (**Figure 2A**), WBC (**Figure 2B**), HGB (**Figure 2C**) and HCT (**Figure 2D**) levels were low or absent in almost all samples. After one day, a subset of samples had residual levels of RBC (Streck: 4/11; Sarstedt: 5/11). At *5 Days*, while absent in Sarstedt plasma, proportionally more Streck plasma samples (4/6) had residual RBC levels. WBC levels were more constantly detected across all samples (Streck (4/6); Sarstedt (3/6); **Supplementary Table 5**), while HGB and HCT remained undetected. At *10 Days*, RBC and HCT levels were absent in all samples, and only very few samples were positive for WBC (Streck: 2/7) and HGB (Sarstedt: 2/7) (**Supplementary Table 5**).

In contrast, platelet-associated parameters (PLT (**Figure 2E**) and PCT (**Figure 2F**)) revealed more significant differences between both BCT types. Streck plasma samples had consistently and mostly significantly higher PLT counts (*5 Days*: p = 0.0143; *10 Days*: p = 0.0493; **Supplementary Table 6**) and PCT ratios (*1 Day*: p = 0.00025; *5 Days*: p = 0.00619; *10 Days*: p = 0.0102; **Supplementary Table 6**) compared to Sarstedt plasma samples.

When investigating differences looking into the interaction of time and BCTs (not shown), many parameters appear to change significantly over time between either the same or the opposite BCT (**Supplementary Table 7**). These results must be taken with caution, as the differential significance might arise from a technical limit of detection.

These findings suggest that plasma from Sarstedt BCTs yield overall lower residual cellular content, indicating more efficient cell removal and stabilization. Temporal trends were observed in both BCT types, with overall lower cell-associated levels at *10 Days* compared to earlier time points, suggesting progressive containment over time or improved clearance of cellular particles upon centrifugation.

### Streck BCTs preserve cfDNA integrity and yield over time more effectively than Sarstedt BCTs, with isolation method playing a secondary role

Aside from the effects of BCTs and prolonged standing times, the cfDNA isolation method further influences cfDNA yield, purity, and fragment integrity. Two established cfDNA isolation kits (MagMAX™ and QIAamp®) were evaluated in combination with two BCT types (Streck and Sarstedt) across multiple time points to identify the most suitable workflow for high-standard molecular diagnostics (**Supplementary Table 6**). To comprehensively assess cfDNA integrity, multiple complementary assays were applied.

Total double-stranded DNA (dsDNA) concentrations were quantified using the Qubit dsDNA High Sensitivity assay. While concentrations were comparable after 1 day, Sarstedt-based workflows yielded in part significantly (StreMax vs SarQia, p = 0.00648) lower DNA concentrations compared to Streck-based workflows after 5 days (**Figure 3A**, **Supplementary Table 8** & **9**). Absolute concentration changes relative to day 1 (Δ) were also greater for Sarstedt-based workflows (**Supplementary Table 8**). At *10 Days*, total DNA concentrations increased significantly (**Supplementary Table 10**) across almost all workflows but with markedly different magnitudes (**Figure 3A, Supplementary Table 8**). Sarstedt-based workflows showed higher median concentrations and substantially increased variance (**Figure 3A**, **Supplementary Table 8** & **9**). Notably, Sarstedt-based workflows exhibited a nine- to ten-fold increase in median DNA concentration at *10 Days*, whereas Streck-based workflows showed more moderate increases (**Supplementary Table 9**).

**Figure 3.**
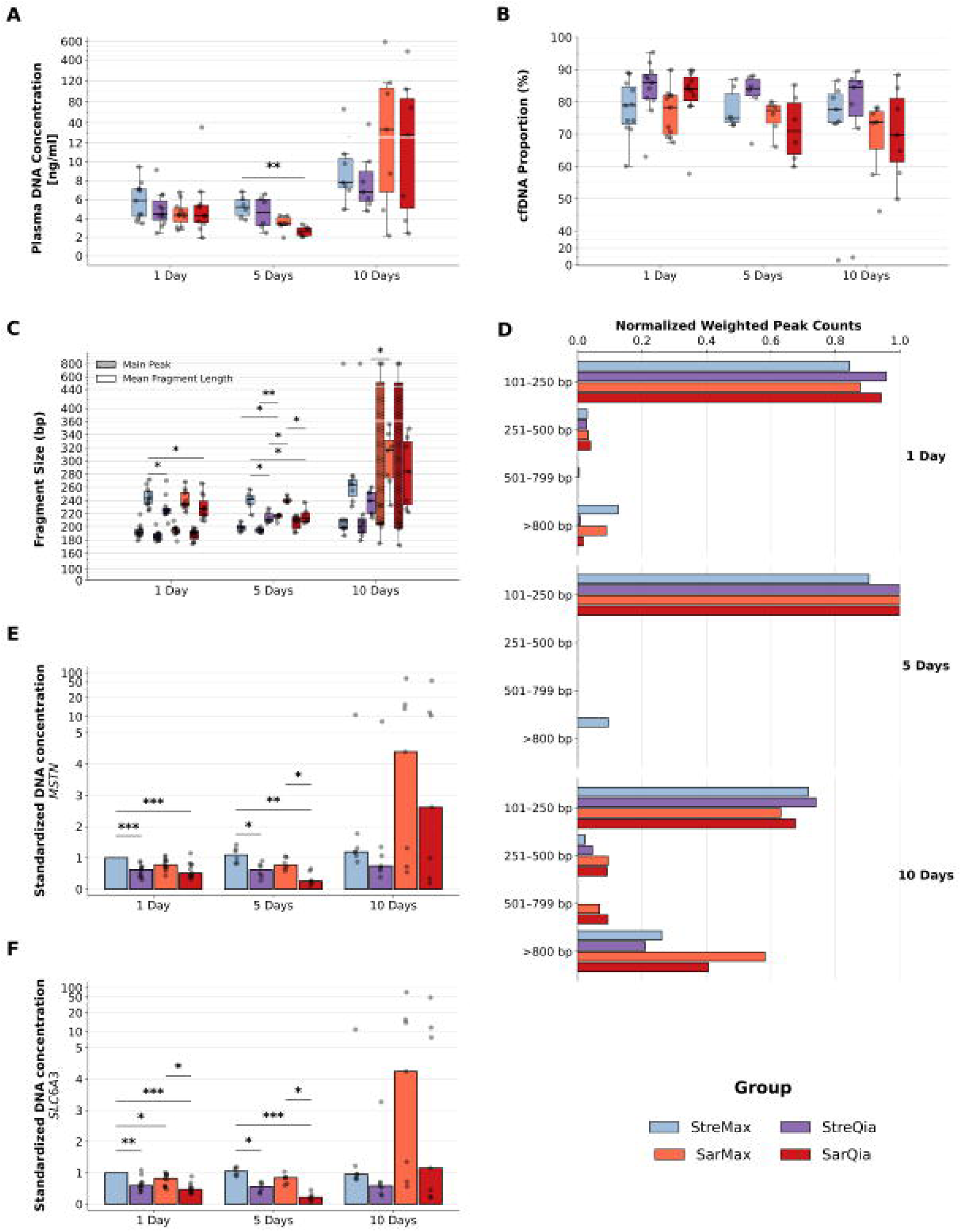
Quantitative characterization of cfDNA yield, integrity, and stability across blood collection tubes and cfDNA isolation workflows during delayed processing. A) Boxplot illustrating total dsDNA concentrations with pronounced increases and variability in Sarstedt-based workflows over time. B) Boxplot illustrating cfDNA ratios stability in Streck-based workflows compared to Sarstedt-based workflows. C) Grouped boxplot illustrating how the main peak (hashed) varies compared to the average fragment lengths (blank) between groups over time. Streck-based workflows exhibited a greater stability compared to Sarstedt-based workflows. D) Time-faceted bar plots illustrating the proportion weighted fragment size distributions for the different workflows. There is an overall accumulation of larger fragments after 10 days, which is again more pronounced in Sarstedt-based workflows. E-F) Bar plots illustrating qPCR results of the single-copy genes (E) MSTN and (F) SLC6A3. StreMax at 1 Day is the reference group to normalize the data against. For both genes, Streck-based workflows exhibited constant concentrations over time, while Sarstedt-based workflows displayed marked pronounced increases and variances after 10 Days. *, p ≤ 0.05; **, p ≤ 0.01; ***, p ≤ 0.001.

Because Qubit quantifies total DNA without discriminating cfDNA from high-molecular-weight DNA, fragment analysis was performed to assess cfDNA ratios and size distributions (**Figure 3B–D, Supplementary Table 8**). Although no overall significant differences were detected (**Figure 3B**, **Supplementary Table 8** & **9**), Streck-based workflows consistently maintained cfDNA ratios above 74.9% (74.92%–78.89%), with StreQia even exceeding 84.1% (84.15%–85.95%) across all time points (**Figure 3B**). Sarstedt-based workflows showed greater variability. While both Sarstedt-based workflows performed comparably at *1 Day*, SarMax maintained high cfDNA ratios over time, while SarQia declined to 70.98% at *5 Days* (Δ = −13.1%) and 69.67% at *10 Days* (Δ = −14.41%) (**Figure 3B**, **Supplementary Table 8**). SarQia also exhibited markedly increased inner-group variance at *5* (IQR = 15.83%) and *10 Days* (IQR = 19.70%) (**Supplementary Table 8**). Using the cfDNA ratio as normalization factor to estimate actual cfDNA specific concentrations, results in drops by 14.05% for StreQia at *1 Day* up to 30.33% for SarQia at *10 Days* (**Supplementary Figure 3**) were observed. Taken together, the proportion of HMW/gDNA is small, but increases greatly in Sarstedt-based workflows at *10 Days* (**Supplementary Figure 4**). For each timepoint, the lowest HMW/gDNA ratios were observed in Qiagen combined workflows (**Supplementary Figure 4**), reflecting their high cfDNA ratios.

Fragment length (FL) profiling further highlighted workflow-dependent differences (**Figure 3C**). At *1 Day*, all workflows showed similar main peaks (185–192 bp) (**Supplementary Table 8**). At *5 Days*, Streck-based workflows remained stable, whereas Sarstedt-based workflows displayed pronounced increases in main peak size (SarMax: Δ = +25.5 bp; SarQia: Δ = +19.0 bp), with significant differences between SarMax and both StreMax (p = 0.0297) and StreQia (p = 0.0047) (**Figure 3C**, **Supplementary Table 9**). Although no significant differences were observed at *10 Days*, Sarstedt workflows again showed greater variability.

Mean FL differences were primarily driven by the isolation method. QIAamp®-based workflows consistently yielded, and often significantly, shorter mean FLs than MagMAX®-based workflows (**Figure 3C**, **Supplementary Tables 8–10**). Between *1 Day* and *5 Days*, QIAamp®-based workflows showed larger decreases in mean FL (Δ = −14.0 bp for both) compared to MagMAX® workflows (StreMax: Δ = −1.5 bp; SarMax: Δ = +3.5 bp). At *10 Days*, mean FL increased across all workflows, with Sarstedt-based workflows again showing significant increases (**Supplementary Table 10**), larger deviations from baseline and greater variance than Streck-based workflows (**Supplementary Table 8**). Correspondingly, differences between mean FL and main peak (Δ_MFL–MP_) were low after 1 and 5 days but increased at *10 Days*, particularly in Sarstedt workflows (**Supplementary Table 11**).

Aggregation of fragment peaks into biologically defined size classes revealed mononucleosomal dominance across all workflows at *1 Day* and *5 Days* (**Figure 3D**). At *1 Day*, StreQia showed the highest mononucleosomal fraction, whereas StreMax exhibited the greatest contribution of >800 bp fragments. By day 5, >800 bp fragments were detectable only in StreMax. At *10 Days*, all workflows showed an increased fraction of longer fragments, including low-level di-nucleosomal peaks. Tri-nucleosomal fragments and a pronounced accumulation of >800 bp DNA fragments were observed exclusively in Sarstedt-based workflows, indicating progressive loss of cfDNA integrity.

Finally, cfDNA concentrations were reassessed by semi-absolute qPCR targeting the single-copy genes *SLC6A3* and *MSTN* (**Figure 3E & 3F, Supplementary Table 8**). QIAamp®-based workflows consistently, and most often significantly, produced lower signals. At 1 day, all workflows yielded significantly lower values than the reference StreMax. After 5 days, StreMax, StreQia, and SarMax remained stable, whereas SarQia showed a pronounced decline. At *10 Days*, Streck-based workflows exhibited minimal changes, while Sarstedt-based workflows showed strong, time-dependent increases in apparent DNA concentration with high variability (**Supplementary Table 8**), consistent with genomic DNA release during prolonged storage.

Across all orthogonal analyses, Streck-based workflows demonstrated superior stability of cfDNA yield, purity, and fragment integrity over prolonged storage. Streck tubes maintained physiologically plausible DNA concentrations, consistently high cfDNA ratios, stable fragment-length profiles, and minimal accumulation of high-molecular-weight DNA for up to 10 days. In contrast, Sarstedt-based workflows showed pronounced temporal instability, characterized by increasing DNA concentrations, declining cfDNA ratios, elevated fragment-length variability, and accumulation of >800 bp fragments indicative of leukocyte degradation and genomic DNA contamination. Among extraction methods, MagMAX™ generally yielded higher and more stable total DNA concentrations, while QIAamp® workflows produced shorter fragment lengths but were more sensitive to storage-related degradation, particularly when combined with Sarstedt BCTs.

## Discussion

Circulating cell-free DNA from liquid biopsies provides disease-relevant genetic and epigenetic information applicable to a wide range of clinical scenarios (1–5). Given that cfDNA is typically present at low concentrations, maintaining its integrity during sample collection, storage and processing is critical for reliable downstream analyses. The growing clinical interest in non-invasive liquid biopsy approaches therefore demands optimized, standardized and robust workflows to consistently maximize cfDNA yield and integrity while minimizing contamination with high molecular weight contaminants (10,19,20). The BCT type, storage duration until plasma isolation, and cfDNA isolation procedure are among the most crucial, influential factors in that regard (3).

This study shows clear differences between BCTs with respect to residual cellular content and long-term cfDNA stability. Prior to plasma isolation, all parameters already had low and over time decreasing levels reflecting the stabilizing capabilities of both BCTs. Most importantly WBC counts and HGB, indicators for HMW contamination, were low or even absent, exceeding results seen in K_2_-EDTA blood plasma (21,22). The isolated Sarstedt plasma was characterized by overall lower residual cellular contamination, most prominently reflected by significantly lower platelet counts and plateletcrit values compared to isolated Streck plasma. Other residual contaminants were largely absent in plasma from both BCT types.

In contrast, for cfDNA yield and integrity Streck-based workflows resulted in more stable and physiologically plausible DNA levels over time. Although total DNA concentrations increased over time across all workflows, markedly stronger time-dependent increases and higher variability were detected in Sarstedt-based workflows, consistent with progressive HMW/genomic DNA release. This was corroborated by fragment analyses, as high cfDNA ratios, stable fragment-length profiles, and less accumulation of HMW fragments were observed even at *10 Days* in Streck-based workflows, which conflicts with Sarstedt’s white paper results (23). While a different cfDNA isolation kit was used in that study, thus potentially explaining the differences, the results presented here can’t be explained by the isolation kit alone. Even though Qiagen’s MinElute ccfDNA kit slightly outperformed ThermoFisher’s MagMAX cell-free DNA isolation kit in regards of cfDNA ratios and fragment distribution, it underscored in means of cfDNA yield. Overall, the differences were mostly small and unsignificant, which is consistent with most reports comparing bead-based versus silica-column-based isolation methods (24–26).

These differences might arise from the differing stabilizing chemistry mix of the BCTs tested. Both BCTs use formaldehyde (FA) as the main stabilizing agent, however, Streck BCTs contain imidazolidinyl urea (IDU) as FA releasing agent, glycine as a quencher, and EDTA as anti-coagulant, and Sarstedt BCTs rely on methenamine (MA) for FA release and citric acid monohydrate for acidification and anti-coagulation. The absence of glycine as FA quencher in Sarstedt BCTs could promote aggregation and more efficient removal of cell components during processing (27). However, prolonged FA incubation can potentially increase cell membrane rigidity (28) turning cells more susceptible to membrane rupture (29) during mechanical stress, giving rise to increased HMW/gDNA contamination at *10 Days*. Additionally, this might be exacerbated by incrementally higher concentrations of FA due to the more potent FA-releasing agent MA in Sarstedt BCTs as compared to IDU in Streck BCTs (30)]. The initial drop in cfDNA concentrations after 5 days for Sarstedt-based workflows, might be due to the progressive FA crosslinking resulting in protease-resistance (31), further exacerbated in combination with the Qiagen MinElute ccfDNA kit as proteinase K digestion efficacy is reduced at room temperature (32). Another possibility could be the absence of EDTA in Sarstedt BCTs as it has been shown to have DNase-inhibitory-like properties (33). This might allow residual nucleases to further degrade cfDNA over time.

## Conclusion

Taken together, the results from this study show that although more efficient initial cellular clearance was achieved with Sarstedt BCTs, superior long-term consistently robust conservation of cfDNA was provided by Streck BCTs. The difference between BCT type performance was largely independent of the isolation method. The most crucial factor however is time. Even though these specialized tubes are advertised for prolonged high-quality conservation of cfDNA, the results here, in accordance with scientific literature (34), underlines the negative impact of delayed sample processing after 5 days.

## Authors’ recommendations

The main goal of this study was to provide a recommendation for a standard operating procedure regarding the isolation and testing of cfDNA for downstream NGS applications in a clinical molecular diagnostic setting. The authors base their recommendation on robustness and reproducibility. Therefore, they defined the following three requisites:

- All steps can be done at room temperature
- A simplified two-step centrifugation protocol for the isolation of plasma using a standard large bench top centrifuge
- cfDNA isolations must be performed according to manufacturer’s instructions

Based on these constants and presented results, the authors conclude that the most optimal combination in BCT, storage delay and isolation kit is:

- Streck BCTs
- Flexible plasma isolation period of one to 5 days
- Either one of the cfDNA isolation kits

## Limitation of the study

This study was designed for clinical settings based on the clinical and laboratory setup at IMGAG in Tübingen. The proposed protocol does not include and investigate harsh constraints present in other settings. Here, most significantly, it was tested and shown that extended standing times influence cfDNA yield and quality, whereas temperature, another impactful variable, was not investigated. A parameter that should be included in a follow-up study. Furthermore, the study concluded that Streck BCTs remains the most suitable collection tube, while the isolation method only plays a secondary role. However, many other collection tubes and isolation kits are available and could have been included. Particularly EDTA-based BCTs are the most used tubes in clinical, biobank and research settings. Testing these specialized tubes would have benefitted from a baseline comparison to K_2_- or K_3_-EDTA tubes. On top, this study did not investigate different centrifugation protocols, which was intentionally another constant to fit the laboratory setup at IMGAG. Furthermore, investigating long time storage at −20°C or −80°C, especially when considering including biobank samples in a study, could be a meaningful next step in refining and diversifying cfDNA protocols.

## Author contribution

MA and KJS performed experiments and analyzed the data; JB performed experiments; ML collected blood samples; LH and CS recruited the patients; MA, SO, CS and KJS wrote and edited the manuscript; CS and KJS designed the study.

## Data Availability

All data produced in the present study are available upon reasonable request to the authors

## Acknowledgements

The authors would like to thank André Koch for his support regarding the complete blood count assay using the Sysmex as well as Nicolas Casedei and his team at the NCTT in Tübingen for their support in the qPCR analyses.

## Supplementary Figures Legends

**Supplementary Figure 1 - Sample collection design.**

Venous blood was collected from healthy participants into two Cell-Free DNA BCT® tubes (Streck; yellow cap) and two S-Monovette® cfDNA Exact tubes (Sarstedt; pink cap) per donor. Samples were assigned to experimental groups based on standing duration prior to plasma isolation and downstream processing: short-term standing time and long-term standing time at room temperature. For both groups, plasma isolation and Sysmex hematology analysis were performed after 24 h (1 day, D01) of standing time. In the short-term standing group, plasma was additionally isolated from a second set of Streck and Sarstedt tubes after 120 h (5 days; D05), whereas in the long-term storage group, plasma isolation was performed after 240 h (10 days; D10). Plasma isolated at each time point was subsequently processed for cell-free DNA (cfDNA) isolations.

**Supplementary Figure 2 - Hematological assessment of whole blood stored in Streck or Sarstedt tubes prior to plasma isolation.**

Whole blood collected in Streck Cell-Free DNA BCT® or Sarstedt S-Monovette® cfDNA Exact tubes was analyzed using a Sysmex XP-300 hematology analyzer after 1 day (D01; n = 11), 5 days (D05; n = 6), and 10 days (D10; n = 7) of storage at room temperature. A) Red Blood Cells (RBCs) were predominantly detected in Streck tubes after 1 day (0.04 ×10D/µL) and were significantly lower in Sarstedt tubes (p ≤ 0.05) as well as at later time points. B) White blood cell (WBC) counts reached their highest values in Sarstedt tubes after 1 day (0.3 ×10³/µL), whereas all other conditions remained below 0.1 ×10³/µL. C) Hemoglobin (HGB) was not detected at any time point in either tube type. D) Hematocrit (HCT) was measurable only after 1 day in both tube types (0.1%; p ≤ 0.05). E) Platelets (PLT) were highest after 1 day in both Streck (424 ×10³/µL) and Sarstedt tubes (414 ×10³/µL) and decreased markedly at later time points. F) Plateletcrit (PCT) exhibited a comparable pattern, with maximal values after 1 day (0.39% in Streck tubes and 0.29% in Sarstedt tubes). Data were presented as medians. Statistics were performed using the Mann–Whitney U test. Significance was defined as follows: p > 0.05, not significant (ns); p ≤ 0.05, *; p ≤ 0.01, **; p ≤ 0.001, ***; p ≤ 0.0001, ****

**Supplementary Figure 3 - Calculated cell-free DNA (cfDNA) concentrations from Qubit and TapeStation measurements.**

Plasma obtained after 1, 5, or 10 days of storage at room temperature in Streck Cell-Free DNA BCT® or Sarstedt S-Monovette® cfDNA Exact tubes was processed using either the MagMAX™ or QIAamp® cfDNA isolation kits, resulting in four analytical workflows: StreMax, StreQia, SarMax, and SarQia. After 1 day, cfDNA concentrations were highest for StreMax (4.20 ng/mL), followed by SarQia (3.81 ng/mL), StreQia (3.61 ng/mL), and SarMax (3.15 ng/mL). After 5 days, cfDNA concentrations remained largely stable for StreQia (3.78 ng/mL; Δ = +0.18 ng/mL), whereas a pronounced decrease was observed for SarQia (1.92 ng/mL; Δ = −1.89 ng/mL), which was significantly lower compared to StreMax (p ≤ 0.01) and StreQia (p ≤ 0.05). After 10 days, Sarstedt-based workflows marked increases with high variances in cfDNA concentrations, with SarMax reaching 20.30 ng/mL (Δ = +17.16 ng/mL) and SarQia 13.72 ng/mL (Δ = +9.91 ng/mL). In contrast, Streck-based workflows remained comparatively stable over time, with StreMax at 5.67 ng/mL (Δ = +1.47 ng/mL) and StreQia at 5.45 ng/mL (Δ = +1.84 ng/mL). Data are presented as median with interquartile ranges and changes relative to Day 1 per group (Δ). Statistical analyses were performed using Kruskal–Wallis tests with Dunn’s post hoc correction and Benjamini–Hochberg adjustment, and interpreted as: p > 0.05 = not significant (ns), p ≤ 0.05 = *, p ≤ 0.01 = **, p ≤ 0.001 = ***, p ≤ 0.0001 = ****.

**Supplementary Figure 4 - Plasma DNA Concentration split by fragment length into short (50-800 bp) and long (>800 bp) fractions.**

Plasma DNA concentrations were subdivided into short fragments (50-800 bp), representing cell-free DNA (cfDNA), and long fragments (>800 bp), representing genomic DNA (gDNA), based on their characteristic size ranges. Plasma obtained after 1, 5, or 10 days of storage at room temperature in Streck Cell-Free DNA BCT® or Sarstedt S-Monovette® cfDNA Exact tubes was processed using either the MagMAX™ or QIAamp® cfDNA isolation kits, generating four analytical workflows: StreMax, StreQia, SarMax, and SarQia. After 1 day, the highest cfDNA concentration was observed for StreMax (4.20 ng/mL), which also exhibited the highest gDNA fraction (1.13 ng/mL). In contrast, SarMax showed the lowest cfDNA concentration (3.51 ng/mL) while maintaining a relatively high gDNA fraction (0.90 ng/mL). StreQia and SarQia yielded moderate cfDNA and gDNA concentrations. After 5 days, MagMAX™-based workflows (StreMax and SarMax) demonstrated more pronounced decreases in both cfDNA and gDNA concentrations, whereas StreQia showed a slight increase in cfDNA concentration to 3.78 ng/mL (Δ = +0.18 ng/mL). SarQia exhibited a marked reduction in cfDNA concentration to 1.92 ng/mL (Δ = −1.89 ng/mL). After 10 days, Streck-based conditions showed only moderate increases in both cfDNA and gDNA concentrations, whereas Sarstedt-based conditions (SarMax and SarQia) induced high increases in both, cfDNA and gDNA content, indicating pronounced DNA release over prolonged storage.

## Supplementary Tables Legends

**Supplementary Table 1 - Donor metadata. Supplementary Table 2 - Study exclusion criteria**

**Supplementary Table 3 - All measured blood count parameters using the Sysmex XP-300 hematology analyzer before and after plasma isolation.**

**Supplementary Table 4 - A) Target gene primer pairs and B) Thermal cycling conditions for amplification of SLC6A3 and MSTN via quantitative PCR using the LightCycler 480 system.**

**Supplementary Table 5 - Summary table for selected Sysmex variables prior and post plasma isolation**

**Supplementary Table 6 - Sysmex statistical significance summary per timepoint per variable**

**Supplementary Table 7 - Sysmex statistical significance summary across timepoints and BCTs per variable**

**Supplementary Table 8 - Summary table for cfDNA yield and quality variables**

**Supplementary Table 9 - cfDNA variables statistical significance summary between workflows per timepoints**

**Supplementary Table 10 - cfDNA variables statistical significance summary per workflow across timepoints**

**Supplementary Table 11 - Differences between mean FL and Main Peaks and statistical outcomes**

## List of abbreviations

cfDNA: Cell-free DNA
StreMax: Streck BCTs combined with MagMAX kit
StreQia: Streck BCTs combined with Qiagen kit
SarMax: Sarstedt BCTs combined with MagMAX kit
SarQia: Sarstedt BCTs combined with Qiagen kit
BCT: Blood collection tube
HMW: High molecular weight
RBC: Red blood cells
WBC: White blood cells
HGB: Haemoglobin
HCT: Haematocrit
PLT: Platelet
PCT: Plateletcrit
FL: Fragment length
Δ: Delta referring to results from *1 Day*
FA: Formaldehyde
IDU: Imidazolidinyl urea
MA: Methenamine
gDNA: Genomic DNA
bp: Base pair

