## Supplementary Figures for "Towards a Robust cell-free DNA Isolation Protocol for NGS Applications in a Clinical Molecular Diagnostics Setting"

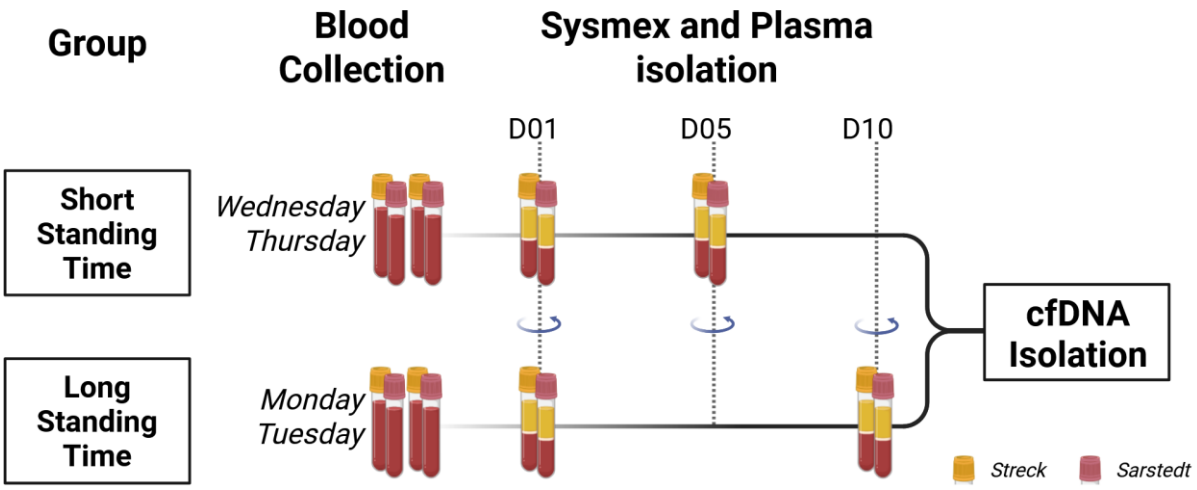


*Supplementary Figure 1: Sample collection design.*

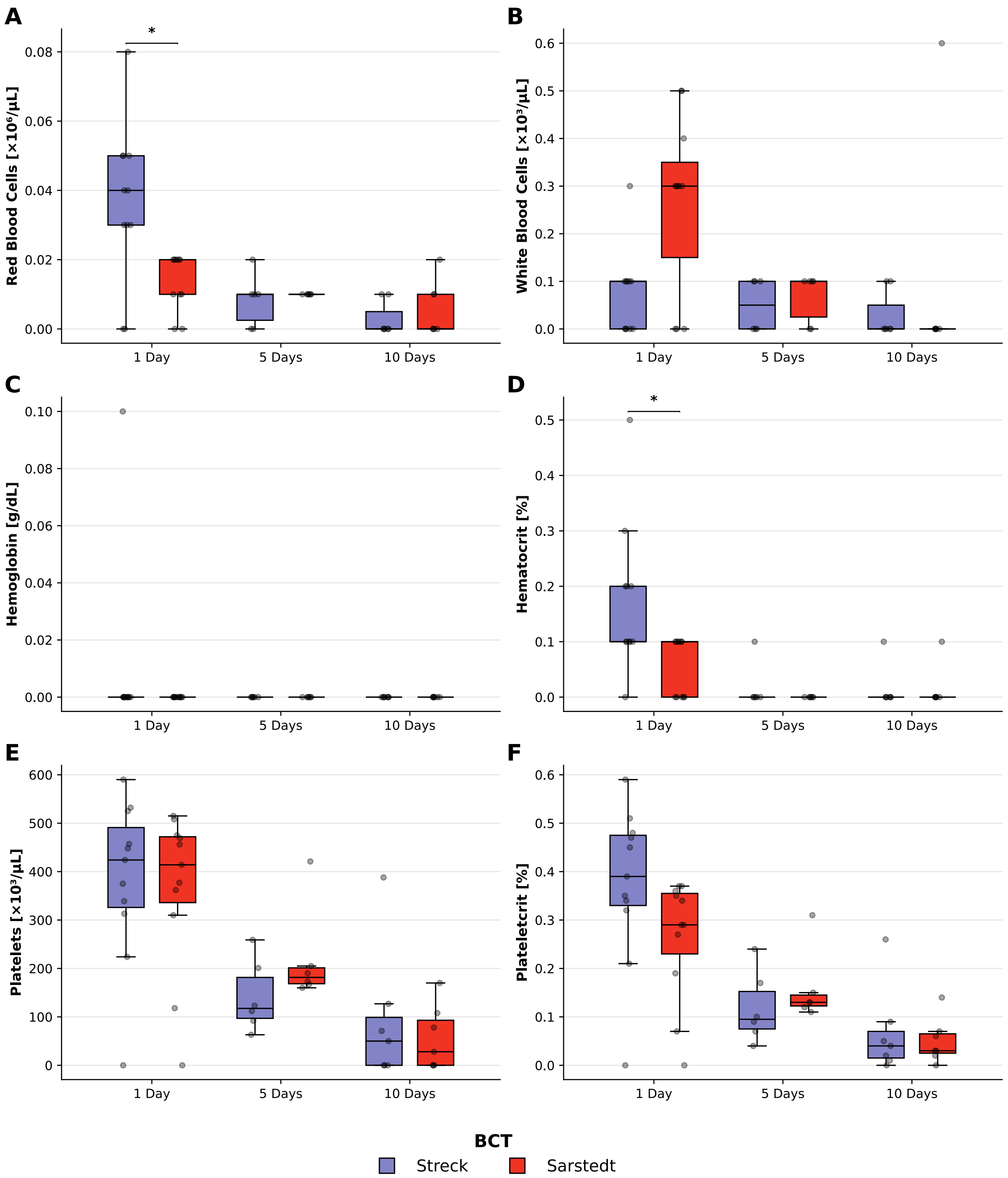


***Supplementary Figure 2: Hematological assessment of whole blood stored in Streck or Sarstedt tubes prior to plasma isolation.***

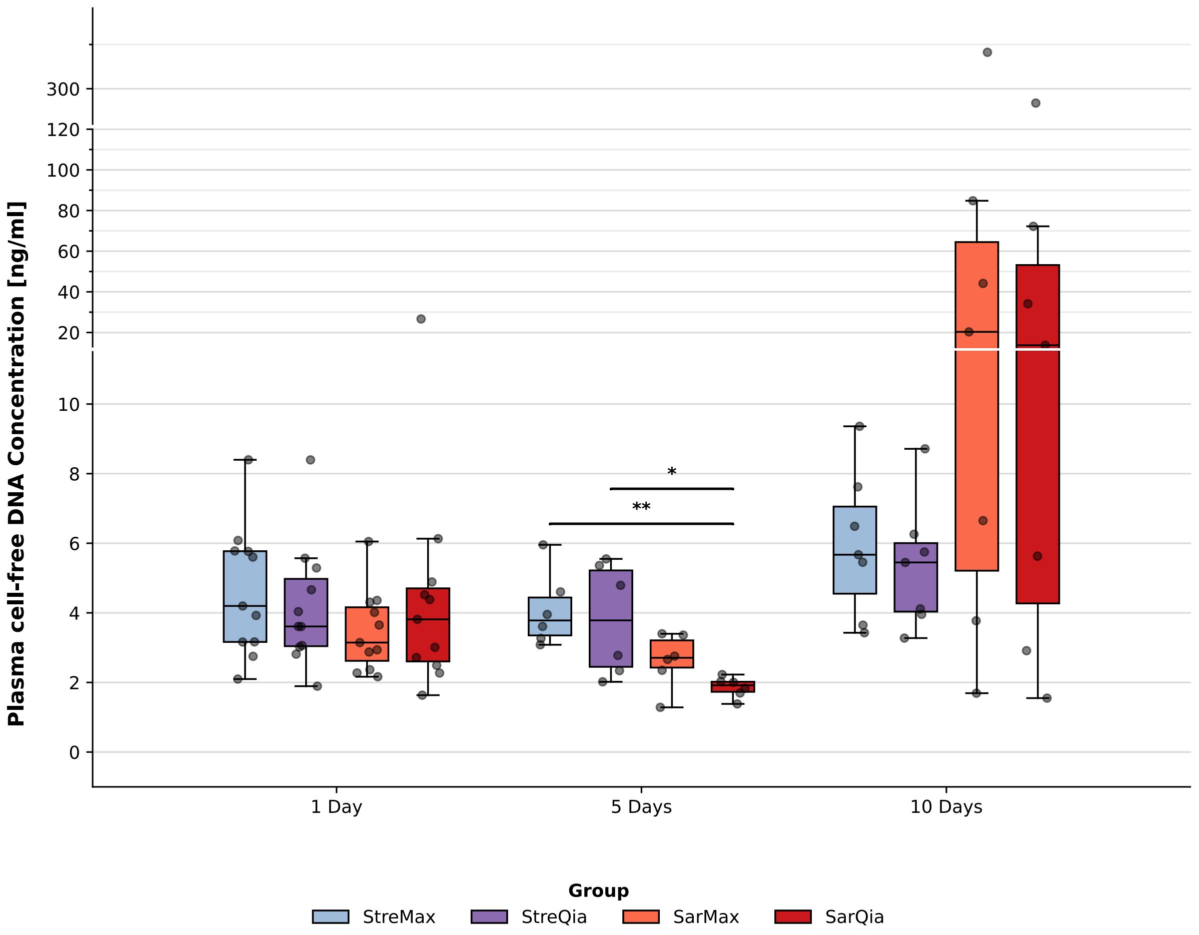


***Supplementary Figure 3: Calculated cell-free DNA (cfDNA) concentrations from Qubit and TapeStation measurements.***

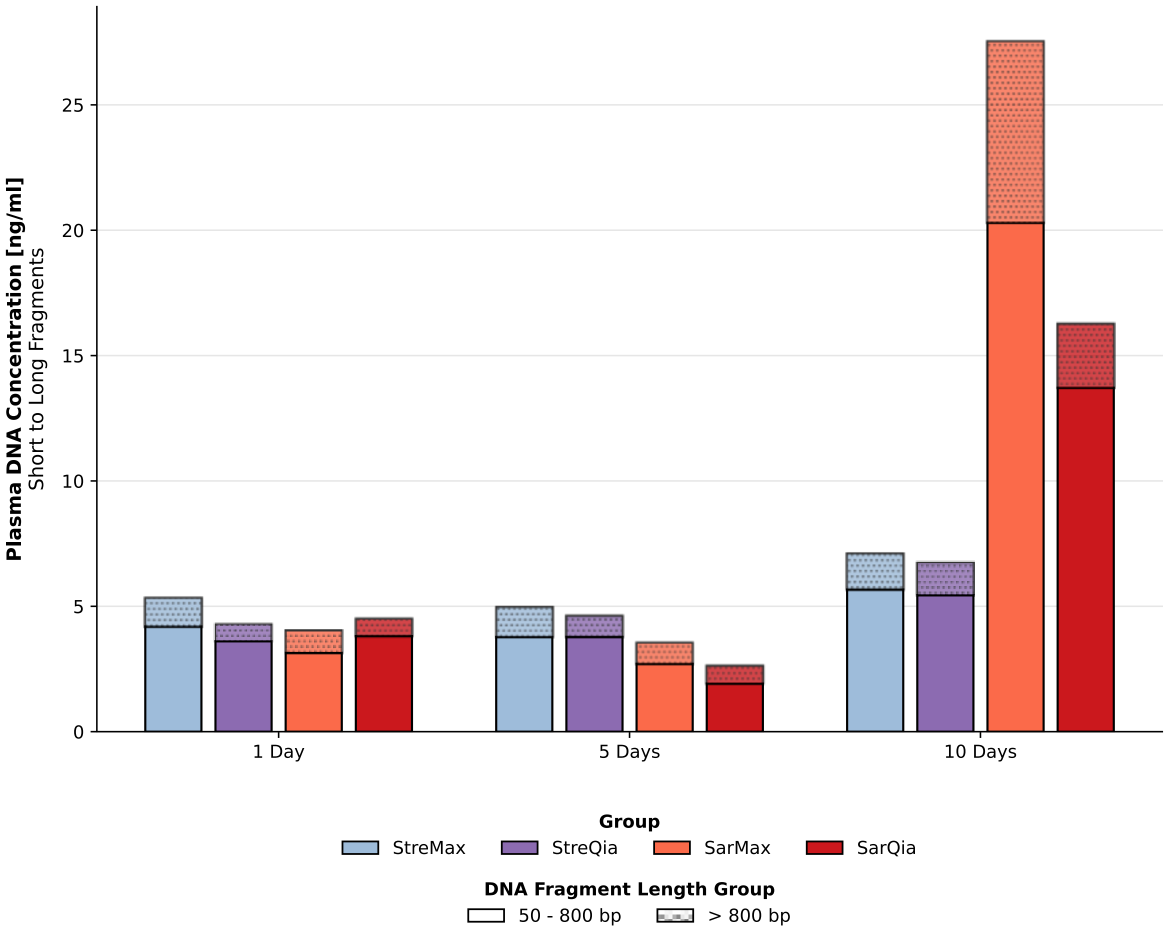


***Supplementary Figure 4: Plasma DNA Concentration split by fragment length into short (50-800 bp) and long (>800 bp) fractions.***
